# Prenatal Acetaminophen, Adverse Birth Outcomes, and ADHD: Mediation Analysis in a Prospective Cohort

**DOI:** 10.1101/2021.06.08.21258568

**Authors:** Brennan H. Baker, Heather H. Burris, Tessa R. Bloomquist, Amélie Boivin, Virginie Gillet, Annie Larouche, Jean-Philippe Bellenger, Jean-Charles Pasquier, Andrea A. Baccarelli, Larissa Takser

## Abstract

**Background:** Although we previously reported an association of prenatal acetaminophen exposure with more than two-fold increased odds of child ADHD, it is unknown whether prenatal acetaminophen is associated with adverse birth outcomes, and if birth outcomes may mediate the association of prenatal acetaminophen with ADHD.

**Methods:** This birth cohort from Sherbrooke, Québec, Canada, included 773 live births. Mothers with no thyroid disease enrolled at their first prenatal care visit or delivery. Acetaminophen was measured in meconium for 393 children at delivery. Physician diagnosis of ADHD was determined when children were 6-7 years old. We first tested associations of prenatal acetaminophen with birthweight, preterm birth, gestational age, and small and large for gestational age. Then we assessed whether these birth outcomes mediate the association of prenatal acetaminophen with ADHD. We imputed missing data via multiple imputation and used inverse probability weighting to account for confounding and selection bias.

**Results:** Acetaminophen was detected in 222 meconium samples (56.5%). Prenatal acetaminophen exposure was associated with decreased birthweight by 136 grams (β = -136; 95%CI [-229, - 43]), 20% increased weekly hazard of delivery (hazard ratio = 1.20; 95%CI [1.00, 1.43]), and over 60% decreased odds of being born large for gestational age (odds ratio = 0.38; 95%CI [0.20, 0.75]). There was no evidence for adverse birth outcomes mediating the association of prenatal acetaminophen with child ADHD.

**Conclusions:** Although unobserved confounding and confounding by indication are possible, these results warrant further investigation into adverse perinatal effects of prenatal acetaminophen exposure.

## Introduction

Acetaminophen (also known as paracetamol) is the only analgesic recommended by doctors for pregnant women, as prenatal exposure to other drugs commonly used to treat pain and fever such as aspirin and non-steroidal anti-inflammatory drugs (e.g. ibuprofen, indomethacin) have previously been associated with birth defects, premature ductus arteriosus closure, and miscarriage.^1-8^ Accordingly, acetaminophen is the most commonly used over-the-counter pain medication taken during pregnancy, with use reported by over half of pregnant women in many populations.^9,10^ During the last two decades, however, research from a multitude of diverse birth cohort studies has revealed consistent associations of prenatal acetaminophen exposure with adverse childhood outcomes including asthma, attention deficit hyperactivity disorder (ADHD), and autism.^11-13^ The severity of these adverse outcomes combined with such high rates of prenatal acetaminophen exposure makes further research into the safety of use during pregnancy an urgent public health priority.

One possibility is that the development of childhood disorders associated with prenatal acetaminophen exposure may be mediated via adverse birth outcomes, such as reduced birth weight and preterm birth. For instance, birth cohort studies have shown associations of pre-pregnancy and prenatal acetaminophen exposure with low birthweight^14^ and preterm birth^10^ respectively, and large meta-analyses have shown associations of low birth weight and preterm birth with asthma,^15,16^ ADHD,^17,18^ and autism.^19,20^

The limited number of studies reporting associations of prenatal acetaminophen with birth outcomes may be a consequence of inaccurate exposure assessment. To the best of our knowledge, all but two cohort studies investigating the effects of prenatal acetaminophen exposure on children’s health have relied on mothers to self-report their acetaminophen use during pregnancy.^21,22^ Furthermore, the only studies to show associations of prenatal acetaminophen with birth outcomes administered questionnaires during pregnancy and postpartum to assess maternal acetaminophen use.^10,14^ Consequently, adverse birth outcomes could have influenced maternal responses in the postpartum interviews; when outcomes are suboptimal, women might be more likely to recall any potential explanatory behavior.^23-25^ Self-reported exposure assessment could also result in misclassification bias towards the null.

The possibility of misclassification bias can be eliminated by measuring prenatal acetaminophen exposure in a biological sample rather than relying on maternal self-report. Measuring chemicals in meconium, the first feces of newborn infants, has proven to be an effective, non-invasive method to assess cumulative prenatal exposures.^22,26,27^ Chemicals in meconium are known to have passed through the fetus and into the fetal intestinal tract,^28-31^ making meconium an ideal substrate for measuring *in utero* exposures. Furthermore, meconium measurements reflect cumulative exposures during the 2^nd^ and 3^rd^ trimesters of pregnancy, as xenobiotics and their metabolites are deposited in meconium throughout that period.^28-31^ Approximately 12% of all deliveries show evidence of meconium stained amniotic fluid, indicating that meconium was passed *in utero*.^32^ Whether meconium is passed *in utero* is likely non-random: risk factors for meconium stained amniotic fluid include advanced gestational age and prolonged labor.^33^ It is therefore important to account for selection bias when relying on exposures measured in this substrate. The primary aim of this study was to evaluate the association of acetaminophen measured in meconium with birth outcomes and pregnancy complications using inverse probability weighting methods to account for confounding and selection bias. The secondary aim was to explore the hypothesis that adverse birth outcomes may link prenatal acetaminophen with ADHD using causal mediation methods.

## Methods

### Study population

This analysis was conducted in the GESTation and the Environment (GESTE) cohort in Sherbrooke, Quebec, Canada. The cohort was initially designed to examine the effects of environmental contaminants on endocrine disruption. Women age ≥18 years without chronic medical conditions and with no known thyroid disease enrolled at the Research Center of the CHUS (Centre Hospitalier Universitaire de Sherbrooke) from September 25, 2007, to September 10, 2009, at their first prenatal care visit or delivery. Among the 800 women recruited, 37 were excluded due to loss to follow up or miscarriage and 10 gave birth to twins, resulting in 773 live births (Figure 1). All study protocols were approved by the institutional review boards of the University of Sherbrooke and Columbia University.

**Figure 1.**
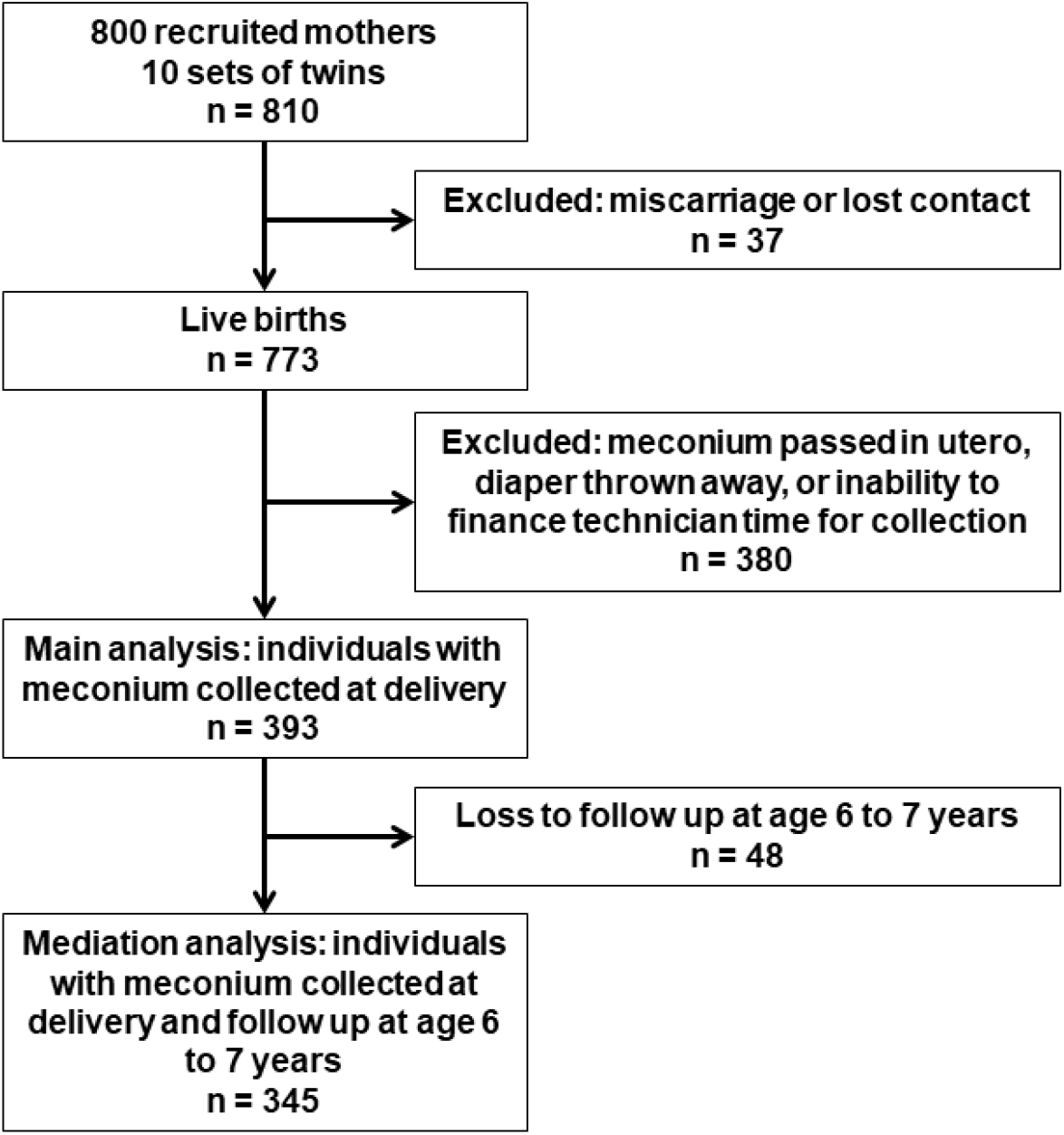
Legend: GESTation and the Environment (GESTE) cohort flowchart

### Exposure assessment

Meconium was collected from diapers of infants after delivery and stored at -80 °C until analysis. Acetaminophen was extracted from < 120 mg meconium and analyzed with ultraperformance liquid chromatography mass spectrometry following the methods described elsewhere.^26^ Among all 773 live births, the eligible study sample was 393 individuals for whom meconium was collected at delivery (Figure 1). Meconium was not always collected, for instance if the infant passed it in utero, if the diaper was thrown in the trash before the technician was able to collect it, or if we were unable to finance technician time for collecting the sample. These potential sources of selection bias were addressed by weighting on the inverse probability of selection (see statistical analysis for details). Acetaminophen was measured with a recovery of 104% and repeatability of ±15%. The limit of detection (LOD) and limit of quantification (LOQ) were 2 ng/g and 5 ng/g respectively.

### Outcomes

#### Birth outcomes

Data for birthweight and gestational age were obtained from CHUS medical records. Infants were weighed on Scale-tronix pediatrics scale 4802 by the obstetric team. Preterm birth was defined as birth before 37 completed weeks of gestation. Small for gestational age (SGA) and large for gestational age (LGA) were defined as birthweights below the 10^th^ percentile and above the 90^th^ percentile for gestational age, respectively. We assigned SGA and LGA categories and computed birthweight for gestational age z-scores in accordance with the Fenton international growth chart.^34^ Data on pregnancy complications, including gestational diabetes, preeclampsia, and high blood pressure, were obtained from CHUS medical records. Complete birth outcome data were available for all 393 individuals with meconium collected at delivery.

#### ADHD

Data on physician diagnosis of ADHD were obtained at a cohort follow-up when children were 6-7 years old or from medical records. These data were available for 345 individuals among the 393 individual study sample (Figure 1).

### Covariates

Covariate data were obtained from CHUS medical records and questionnaires administered after delivery. Covariates were child sex, familial income, and maternal characteristics including age at delivery, education status (College/University vs. no College/University), pre-pregnancy BMI, smoking during pregnancy (yes/no), and alcohol during pregnancy (yes/no).

### Statistical analysis

Differences in baseline characteristics between 1) individuals with and without prenatal acetaminophen exposure, and 2) individuals with and without meconium collected at delivery were determined using chi-square goodness of fit tests for binary variables and two-sample t-tests for continuous variables. Using linear regression, we estimated associations of meconium acetaminophen detection (yes vs. no) with birthweight in grams and birthweight for gestational age z-score. Using logistic regression, we estimated associations of meconium acetaminophen detection with SGA, LGA, preterm birth, and maternal gestational diabetes, preeclampsia, and high blood pressure. Using cox proportional hazards models with birth as the event and gestational age as the time to event, we estimated the association of meconium acetaminophen detection with the hazard for giving birth. Because birth (the event) occurred for all individuals, increased hazards in these models indicate shorter gestational age (the time to event). Confidence intervals for cox models were calculated with robust standard errors. Coefficients from logistic regressions and cox models were exponentiated into odds ratios and hazard ratios respectively. To account for missing covariate data, all models were employed on 10 datasets imputed using the ‘MICE’ R package.^35^ Estimates and standard errors from imputed datasets were combined using Rubin’s Rule.^36,37^

In addition to unadjusted models, we report models adjusted for the covariates described above. We controlled for covariates by inverse probability of exposure weighting (IPW) using propensity scores.^38-41^ Propensity scores (p, the likelihood of detectable meconium acetaminophen) were estimated using logistic regression models in which exposure (meconium acetaminophen detected vs. not detected) was regressed on the covariates described above. Weights were estimated as 1/p for exposed individuals, and 1/(1 – p) for unexposed individuals. Study sample weighting creates a pseudo-population balanced on measured baseline covariates.^38-40^

We additionally addressed potential selection bias related to meconium collection by weighting models on both the inverse probability of exposure described above and the inverse probability of selection. We first created weights for the probability of selection using logistic regression models in which selection (meconium collected vs. not collected) was regressed on the covariates described above for the entire 810 individual cohort. Then we removed those without meconium data and created weights for the probability of exposure as described above before fitting weighted models of the effect of meconium acetaminophen exposure on birth outcomes. Weighting the 393 individual selected sample on the probability of selection creates a pseudo-population that is comparable on measured covariates to the entire 810 individual cohort.

In this cohort, we have previously shown an association of prenatal acetaminophen with increased odds for ADHD,^22^ and associations of ADHD with adverse birth outcomes including preterm birth and lower birthweight.^27^ Here, we additionally assessed whether birth outcomes mediated the relationship between prenatal acetaminophen exposure and ADHD, in separate models for each birth outcome, using the ‘mediation’ R package,^42^ which implements a quasi-Bayesian Monte Carlo method with 1,000 simulations. To test for exposure-mediator interaction, we modeled interactions between meconium acetaminophen and birth outcomes on ADHD in separate models for each birth outcome. Because we found no significant interaction terms, we assumed no exposure-mediator interaction in the mediation models. We estimated the total and direct effects of prenatal acetaminophen on ADHD, as well as the causal mediation (i.e., natural indirect) effects through each birth outcome. We included the covariates discussed above as terms in the mediation models to control for confounders of the exposure-mediator, exposure-outcome, and mediator-outcome relationships. Because quasi-Bayesian confidence intervals cannot be combined with Rubin’s Rule, we report separate covariate adjusted mediation models for each imputed dataset. Statistical analyses were conducted with R, version 3.5.1.^43^

## Results

Baseline covariates stratified by meconium acetaminophen detection are presented in Table 1. Acetaminophen was detected in the meconium of 222 individuals (56.5%) among the total study sample of 393 (Table 1).

**Table 1:** Characteristics of study sample stratified by detection of acetaminophen in meconium in the GESTation and the Environment (GESTE) cohort (n = 393).

|  | No<br>acetaminophen<br>(N=171) | Acetaminophen<br>(N=222) | Total (N=393) | P value |
| --- | --- | --- | --- | --- |
| **Sex** |  |  |  | 0.968 |
| Female | 82 (48.0%) | 106 (47.7%) | 188 (47.8%) |  |
| Male | 89 (52.0%) | 116 (52.3%) | 205 (52.2%) |  |
| **Maternal age at**<br>**delivery (years)** |  |  |  | 0.986 |
| Mean (SD) | 28.9 (4.7) | 28.9 (4.5) | 28.9 (4.6) |  |
| Range | 18.0 - 43.0 | 19.0 - 41.0 | 18.0 - 43.0 |  |
| **Maternal education** |  |  |  | 0.980 |
| No College or<br>University | 68 (39.8%) | 88 (39.6%) | 156 (39.7%) |  |
| College or<br>University | 103 (60.2%) | 134 (60.4%) | 237 (60.3%) |  |
| **Family income**<br>**(Canadian dollars)** |  |  |  | 0.596 |
| N-Miss | 21 | 18 | 39 |  |
| Mean (SD) | 69,874 (47,968) | 67,235<br>(44,858) | 68,353<br>(46,153) |  |
| Range | 2,600 – 500,000 | 8000 – 450,000 | 2,600 –<br>500,000 |  |
| **Maternal BMI**<br>**(kg/m<sup>2</sup>)** |  |  |  | 0.020 |
| N-Miss | 0 | 1 | 1 |  |
| Mean (SD) | 24.9 (5.2) | 26.2 (5.9) | 25.7 (5.6) |  |
| Range | 17.9 - 45.2 | 17.7 - 49.1 | 17.7 - 49.1 |  |
| **Smoked during**<br>**pregnancy** |  |  |  | 0.927 |
| N-Miss | 8 | 5 | 13 |  |
| No | 141 (86.5%) | 187 (86.2%) | 328 (86.3%) |  |
| Yes | 22 (13.5%) | 30 (13.8%) | 52 (13.7%) |  |
| **Alcohol during**<br>**pregnancy** |  |  |  | 0.161 |
| N-Miss | 8 | 5 | 13 |  |
| No | 120 (73.6%) | 173 (79.7%) | 293 (77.1%) |  |
| Yes | 43 (26.4%) | 44 (20.3%) | 87 (22.9%) |  |

Accounting for all covariates, prenatal acetaminophen exposure was associated with decreased birthweight by over 136 grams (β = -136; 95% CI [-229, -43]) and decreased birthweight for gestational age z-score (β = -0.17; 95% CI [-0.34, 0.00]) (Table 2). Consistent with these associations with decreased birthweight, prenatal acetaminophen was also associated with over 60% decreased odds of LGA (odds ratio [OR] = 0.38; 95% CI [0.20, 0.75]) (Table 2). In addition to their decreased birthweight and lower likelihood of LGA, the mean time of gestation was on average 0.3 weeks shorter among individuals exposed to acetaminophen *in utero* compared to the unexposed group (39.0 vs. 39.3 weeks). Accordingly, prenatal acetaminophen exposure was associated with a 20% increased weekly hazard of delivery (hazard ratio = 1.20; 95% CI [1.00, 1.43]), indicating a higher likelihood for earlier delivery and thus reduced gestational age in exposed individuals (Table 2). Our data suggest that prenatal acetaminophen was not associated with SGA or preterm birth (Table 2).

**Table 2:** Association of meconium acetaminophen with birth outcomes (N=393)

|  | Mean or n (%) |  | Coefficient, Hazard Ratio, or Odds Ratio <sup>a</sup> |  |  |
| --- | --- | --- | --- | --- | --- |
|  | Acetaminophen | No Acetaminophen | Unadjusted | Covariate adjusted <sup>b</sup> | Selection bias adjusted <sup>c</sup> |
| Birthweight (g) | 3338 | 3459 | -121<br>[-213, -28] | -136<br>[-229, -43] | -141<br>[-232, -49] |
| Birthweight for gestational age z-score | -0.129 | 0.000 | -0.13<br>[-0.3, 0.04] | -0.17<br>[-0.34, 0] | -0.18<br>[-0.35, -0.01] |
| Gestational age (weeks) | 39.0 | 39.3 | 1.23<br>[1, 1.5] <sup>d</sup> | 1.2<br>[1, 1.43] <sup>d</sup> | 1.21<br>[1.01, 1.45] <sup>d</sup> |
| Small for gestational age | 15 (6.8) | 13 (7.6) | -0.13<br>[-0.9, 0.64] | -0.08<br>[-0.63, 0.46] | 0.01<br>[-0.37, 0.39] |
| Large for gestational age | 7 (3.2) | 12 (7.0) | 0.43<br>[0.17, 1.12] | 0.38<br>[0.2, 0.75] | 0.36<br>[0.22, 0.58] |
| Preterm birth | 8 (3.6) | 9 (5.3) | 0.67<br>[0.25, 1.78] | 0.7<br>[0.35, 1.41] | 0.62<br>[0.38, 1.02] |

<sup>a</sup> Coefficients from linear regression shown for birthweight and birthweight for gestational age z-score. Odds ratios from logistic regression shown for small for gestational age (SGA), large for gestational age (LGA), and preterm birth. Hazard ratios from cox proportional hazard models shown for gestational age.
<sup>b</sup> Adjusted for covariates by inverse probability of exposure weighting using child sex, familial income, and maternal age, education, BMI, smoking during pregnancy, and alcohol during pregnancy to predict exposure.
<sup>c</sup> Adjusted for covariates by inverse probability of exposure weighting and for selection bias by weighting on the inverse of the probability of selection.
<sup>d</sup> Ratio for instantaneous hazard of delivery.

Mothers of children for whom meconium was collected were significantly older and more educated (Supplemental Table 1), indicating the potential for selection bias. Controlling for selection bias by weighting models to account for these covariate differences, however, did not appreciably impact the estimates of the effects of meconium acetaminophen detection on any birth outcome, suggesting minimal bias related to meconium sampling in this cohort (Table 2).

Meconium acetaminophen was not associated with pregnancy complications including gestational diabetes, preeclampsia, or high blood pressure (Table 3).

**Table 3:** Associations of meconium acetaminophen with pregnancy complications (N=393)

|  | Mean or n (%) |  | Odds Ratio |  |  |
| --- | --- | --- | --- | --- | --- |
|  | Acetaminophen | No Acetaminophen | Unadjusted | Covariate adjusted <sup>a</sup> | Selection bias adjusted <sup>b</sup> |
| Gestational diabetes | 30 (13.5) | 20 (11.7) | 1.18<br>[0.64, 2.16] | 1.05<br>[0.69, 1.6] | 1.02<br>[0.76, 1.37] |
| Preeclampsia | 3 (1.4) | 2 (1.2) | 1.16<br>[0.19, 7.01] | 1.01<br>[0.29, 3.53] | 0.88<br>[0.36, 2.16] |
| High blood pressure | 19 (8.6) | 11 (6.4) | 1.36<br>[0.63, 2.94] | 1.08<br>[0.64, 1.83] | 1.13<br>[0.77, 1.64] |

<sup>a</sup> Adjusted for covariates by inverse probability of exposure weighting using child sex, familial income, and maternal age, education, BMI, smoking during pregnancy, and alcohol during pregnancy to predict exposure.
<sup>b</sup> Adjusted for covariates by inverse probability of exposure weighting and for selection bias by weighting on the inverse of the probability of selection.

In this cohort, we previously reported an association of prenatal acetaminophen exposure with more than two-fold increased odds of child ADHD.^22^ Consistent with prior work, we found significant direct and total effects of meconium acetaminophen on ADHD in all mediation models (see Table 4 for unadjusted models, see Supplemental Table 2 for adjusted models in 10 datasets imputed for missing covariates). Causal mediation effects, however, were non-significant for all birth outcomes in both unadjusted and adjusted models.

**Table 4:** Analysis of mediation of the association between prenatal acetaminophen and ADHD by birth outcomes (N = 345). ^a^

| Mediator | Total effect on ADHD |  | Direct effect of prenatal acetaminophen |  | Average causal mediation effect |  |
| --- | --- | --- | --- | --- | --- | --- |
|  | Estimate | P value | Estimate | P value | Estimate | P value |
| Birthweight | 0.07 [0.01, 0.14] | 0.028 | 0.07 [0, 0.13] | 0.036 | 0.007 [-0.003, 0.021] | 0.188 |
| Birthweight for gestational age z-score | 0.07 [0.01, 0.13] | 0.024 | 0.07 [0.01, 0.13] | 0.032 | 0.004 [-0.003, 0.015] | 0.314 |
| Gestational age | 0.07 [0.01, 0.13] | 0.032 | 0.07 [0.01, 0.13] | 0.036 | 0.002 [-0.005, 0.011] | 0.612 |
| Small for gestational age | 0.07 [0.01, 0.13] | 0.02 | 0.08 [0.01, 0.14] | 0.018 | -0.007 [-0.028, 0.009] | 0.354 |
| Large for gestational age | 0.07 [0.01, 0.13] | 0.028 | 0.07 [0.01, 0.13] | 0.028 | 0 [-0.005, 0.006] | 0.89 |
| Preterm birth | 0.07 [0.01, 0.13] | 0.028 | 0.07 [0.01, 0.13] | 0.032 | 0 [-0.015, 0.009] | 0.818 |

<sup>a</sup> Unadjusted models shown. See Supplemental Table 2 for adjusted models in 10 datasets imputed for missing covariates.

## Discussion

In this Eastern Canadian birth cohort, detection of acetaminophen in meconium was associated with decreased birthweight, decreased gestational age, and decreased odds of LGA. Although meconium was only collected for approximately half of our eligible births, we found no evidence that selection bias impacted effect estimation: associations of meconium acetaminophen with outcomes were nearly identical in both models adjusted and not adjusted for selection bias. While adverse birth outcomes such as preterm birth and reduced birthweight are known to be associated with ADHD, we found no evidence for mediation by birth outcomes of the association between prenatal acetaminophen exposure and ADHD in this cohort.

A small number of studies have previously shown associations between maternal self-reported acetaminophen use during pregnancy and adverse birth outcomes. In the Danish National Birth Cohort, there was an increased risk of preterm birth among women using acetaminophen during the third trimester of pregnancy, but there were no associations of acetaminophen use with miscarriage, stillbirth, low birth weight, or SGA, or with common preterm birth complications including bronchopulmonary dysplasia, intracranial hemorrhage, retinopathy of prematurity, perinatal infections and anemia of prematurity.^10^ In the Ontario Birth Study, maternal acetaminophen use in the 3 months before pregnancy was associated with low birthweight and increased risk for SGA, but maternal acetaminophen use during pregnancy was not.^14^ However, both of those studies were prone to substantial recall bias, as they assessed maternal acetaminophen use via questionnaires administered during pregnancy and postpartum. When self-report occurs in postpartum interviews, mothers of infants with adverse birth outcomes may rack their brains for an explanation, thereby overreporting exposures.^23-25^ Our study, on the other hand, utilized a direct measurement of prenatal acetaminophen exposure measured in meconium that is unbiased by inaccurate recall.

While the associations of prenatal acetaminophen exposure with adverse birth outcomes found here may be concerning, more studies in a diverse range of cohorts are needed before suggesting a change in clinical practice. Additionally, mechanisms underlying the associations of prenatal acetaminophen exposure with adverse birth outcomes remain unknown. Acetaminophen may inhibit prostacyclin synthesis and thereby promote pre-eclampsia,^44^ which has previously been associated with intrauterine growth restriction and reduced gestational age^.45^

Acetaminophen exposure may also trigger the immune system and upregulate oxidative stress response pathways^46^ that may underlie adverse birth outcomes. A better understating of the mechanisms through which prenatal acetaminophen exposure may affect birth outcomes is needed, not only to better assess causality, but also to serve as potential targets in future intervention studies.

Our study has several strengths. First, the high genetic and sociodemographic homogeneity in the GESTE cohort limits the likelihood of confounding by unknown genetic or sociodemographic factors. Second, the prospective nature of the cohort limits sources of bias common in retrospective designs, including selection and recall bias. Third, we explicitly controlled for known sources of confounding and selection bias. Finally, our measurement of prenatal acetaminophen exposure in meconium eliminates the possibility of recall bias.

This study has limitations. First, while the homogeneity of the GESTE cohort may limit confounding, it also lowers the generalizability of results. Second, this study had a relatively small sample size of just under 400 mother child pairs. Consequently, there were few events for several outcomes, including early prematurity and severe preeclampsia. Third, we lacked the information necessary to control for indications for acetaminophen use, such as chronic pain, fever, and infections during pregnancy. However, when controlling for indications for acetaminophen in the Danish National Birth Cohort study, Rebordosa and colleagues still observed associations of prenatal acetaminophen with increased risk of preterm birth.^10^ However, results from other cohorts may not be generalizable to this Eastern Canadian population. Therefore, confounding by indication remains a possibility. Another possibility is that acetaminophen was more easily detected in the meconium of smaller infants owing to less efficient metabolism. However, acetaminophen pharmacokinetics in the fetus parallels that in the mother, with fetal and maternal acetaminophen reaching comparable levels as early as 30 minutes after maternal administration.^47^ Inverse causality is thus unlikely. Finally, we did not ask women to self-report their use of acetaminophen during pregnancy, so we were unable to correlate acetaminophen intake with levels of acetaminophen in meconium. Future studies are needed to determine the dosage and timing of acetaminophen required to have detectable levels in meconium.

In conclusion, while this study may add evidence that support questioning the safety of acetaminophen use during pregnancy, more work is needed to rule out confounding by indication and to assess generalizability before a change in clinical practice is recommended. Additionally, our data do not support adverse birth outcomes as the pathway through which acetaminophen may affect the risk of ADHD. Thus, further work to delineate the pathophysiology of prenatal acetaminophen and brain development is warranted.

## Data Availability

Data contain protected health information and are thus confidential. Code used for the analyses are available upon request.

