## Supplemental Table 1 for "Prenatal Acetaminophen, Adverse Birth Outcomes, and ADHD: Mediation Analysis in a Prospective Cohort"

Supplemental Table 1: Characteristics of study sample stratified by meconium collection in the GESTation and the Environment (GESTE) cohort (n = 810).

|  | Meconium collected (N=393) | No meconium collected (N=417) | Total (N=810) | p value |
| --- | --- | --- | --- | --- |
| **Sex** |  |  |  | 0.429 |
| N-Miss | 0 | 37 | 37 |  |
| Female | 188 (47.8%) | 171 (45.0%) | 359 (46.4%) |  |
| Male | 205 (52.2%) | 209 (55.0%) | 414 (53.6%) |  |
| **Maternal age at delivery** |  |  |  | 0.004 |
| N-Miss | 0 | 33 | 33 |  |
| Mean (SD) | 28.91 (4.56) | 27.97 (4.62) | 28.45 (4.61) |  |
| Range | 18.00 - 43.00 | 18.00 - 41.00 | 18.00 - 43.00 |  |
| **Maternal education** |  |  |  | < 0.001 |
| No College or University | 156 (39.7%) | 227 (54.4%) | 383 (47.3%) |  |
| College or University | 237 (60.3%) | 190 (45.6%) | 427 (52.7%) |  |
| **Family income (Canadian dollars)** |  |  |  | 0.052 |
| N-Miss | 39 | 93 | 132 |  |
| Mean (SD) | 68353.67 (46153.47) | 62420.68 (30869.48) | 65518.44 (39675.38) |  |
| Range | 2600.00 - 500000.00 | 7000.00 - 180000.00 | 2600.00 - 500000.00 |  |
| **Maternal BMI** |  |  |  | 0.145 |
| N-Miss | 1 | 7 | 8 |  |
| Mean (SD) | 25.67 (5.64) | 25.06 (6.16) | 25.35 (5.91) |  |
| Range | 17.71 - 49.08 | 14.99 - 55.60 | 14.99 - 55.60 |  |
| **Smoked during pregnancy** |  |  |  | 0.163 |
| N-Miss | 13 | 73 | 86 |  |
| No | 328 (86.3%) | 284 (82.6%) | 612 (84.5%) |  |
| Yes | 52 (13.7%) | 60 (17.4%) | 112 (15.5%) |  |
| **Alcohol during pregnancy** |  |  |  | 0.406 |
| N-Miss | 13 | 73 | 86 |  |
| No | 293 (77.1%) | 274 (79.7%) | 567 (78.3%) |  |
| Yes | 87 (22.9%) | 70 (20.3%) | 157 (21.7%) |  |
